# Vaccine hesitancy and reasons for refusing the COVID-19 vaccination among the U.S. public: A cross-sectional survey

**DOI:** 10.1101/2021.02.28.21252610

**Authors:** Ali S. Raja, Joshua D. Niforatos, Nancy Anaya, Joseph Graterol, Robert M. Rodriguez

**Author notes:** Corresponding Author (ASR).

## Abstract

**Importance:** Although widespread vaccination will be the most important cornerstone of the public health response to the COVID-19 pandemic, a critical question remains as to how much of the United States population will accept it.

**Objective:** Determine: 1) rate of COVID-19 vaccine hesitancy in the United States public, 2) patient characteristics associated with hesitancy, 3) reasons for hesitancy, 4) healthcare sites where vaccine acceptors would prefer to be vaccinated.

**Design:** 43-question cross-sectional survey conducted November 17-18, 2020, distributed on Amazon Mechanical Turk, an online labor marketplace where individuals receive a nominal fee (here, $1.80) for anonymously completing tasks.

**Eligible Participants:** United States residents 18-88 years of age, excluding healthcare workers. A total 1,756 volunteer respondents completed the survey (median age 38 years, 53% female).

**Main Outcome Measure:** Multivariable logistic regression modeled the primary outcome of COVID-19 vaccine hesitancy (defined as non-acceptance or being unsure about acceptance of the COVID-19 vaccine) with respondent characteristics.

**Results:** A total 663 respondents (37.8%) were COVID-19 vaccine hesitant (374 [21.3%] non-acceptors and 289 [16.5%] unsure about accepting). Vaccine hesitancy was associated with not receiving influenza vaccination in the past 5 years (odds ratio [OR] 4.07, 95% confidence interval [CI] 3.26-5.07, p<0.01), female gender (OR 2.12, 95%CI 1.70-2.65, p<0.01), Black race (OR 1.54, 95%CI 1.05-2.26, p=0.03), having a high school education or less (OR 1.46, 95%CI 1.03-2.07, p=0.03), and Republican party affiliation (OR 2.41, 95%CI 1.88-3.10, p<0.01). Primary reasons for hesitancy were concerns about side effects, need for more information, and doubts about vaccine efficacy. Preferred sites for vaccination for acceptors were primary doctors’ offices/clinics, pharmacies, and dedicated vaccination locations.

**Conclusions:** In this recent national survey, over one-third of respondents were COVID-19 vaccine hesitant. To increase vaccine acceptance, public health interventions should target vaccine hesitant populations with messaging that addresses their concerns about safety and efficacy.

## INTRODUCTION

The greatest public health crisis of the past century, the COVID-19 pandemic has led to over 1.8 million deaths globally as of January 3, 2021 (1). The three tenets of the public health response to the pandemic remain social distancing, mask wearing, and vaccination (2,3). However, these mitigation measures are only as effective as their broad acceptance and implementation.

Along with research and development of therapeutics, the most anticipated control measures are a series of COVID-19 vaccines, two of which - as of this writing - have received United States (U.S.) Food and Drug Administration emergency use authorizations (4). As COVID-19 vaccination is implemented across the U.S., a critical question remains as to how much of the population will accept it. For COVID-19 vaccination to effectively confer herd immunity, experts agree that at least 60-70% of the population will need to be vaccinated (5). Vaccine hesitancy, a phenomenon which predates the pandemic, has been well studied with other vaccinations, including the influenza and Measles/Mumps/Rubella vaccines. Recent influenza vaccine vaccination hesitancy rates have hovered at approximately 40% (6–9). The traditionally low rates of influenza vaccination in Black, Latinx, and Native American populations are of particular concern since these groups have had disproportionately poor outcomes during the COVID-19 pandemic (9–11). While a recent study found that COVID-19 vaccine hesitancy rates have varied between 26-44% (with rates increasing throughout 2020), the reasons for vaccine refusal in late 2020 have yet to be fully described (12). These reasons are especially relevant as we begin public vaccination programs in early 2021.

With the need for widespread acceptance of COVID-19 vaccination in mind, the objectives of this survey study were to determine: 1) the US population rate of COVID-19 vaccine hesitancy (defined as either non-acceptance or unsure about acceptance of the COVID-19 vaccine), 2) characteristics associated with hesitancy, 3) reasons for hesitancy, and 4) health care sites where respondents would prefer to receive the vaccine.

## MATERIALS AND METHODS

### Study Setting and Population

We distributed this cross-sectional survey from November 17 to November 18, 2020 on Amazon Mechanical Turk (MTurk, https://www.mturk.com), an online labor marketplace in which individuals anonymously complete tasks, including surveys, and in return receive a nominal fee (in this case, $1.80). MTurk is well-validated for behavioral experiments and increasingly used to study healthcare questions, and data from MTurk are considered reliable (13,14). This study was approved by the Institutional Review Board at <redacted for review>.

We recruited U.S. residents between 18 and 88 years of age from MTurk to complete a 43-question survey. Because our goal was to assess vaccine hesitancy in a more medically naïve population, we excluded respondents self-identifying as healthcare workers.

### Survey Instrument

The survey (Supplement) included questions regarding demographic characteristics, health insurance status, healthcare utilization, employment and housing status, and political affiliation. Survey respondents were then asked a series of questions regarding self-reported adherence to different COVID-19 mitigation measures and previous influenza vaccinations. After a short descriptor about the COVID-19 vaccine including the statement that it would likely be provided free of charge, participants were asked, “Would you accept the COVID-19 vaccine when it becomes available?” Respondents who responded that they would accept it were then asked their preferred location to receive a COVID-19 vaccine. The survey also contained quality assurance questions to ensure meaningful responses. Respondents not appropriately responding to these questions were excluded from analyses.

### Primary and Secondary Outcome Measures

The primary outcome measure was COVID-19 vaccine hesitancy - defined as either non-acceptance or being unsure about acceptance of the COVID-19 vaccine. Other outcomes included patient characteristics associated with vaccine hesitancy, reasons for hesitancy, and health care sites where vaccine acceptors would prefer to be vaccinated.

### Statistical Analysis

We coded survey items as continuous, ordinal, or categorical variables in accordance with their survey presentation and report respondent demographics using standard descriptive statistics, e.g., medians and interquartile ranges (IQRs). We transformed the primary outcome of COVID-19 vaccine hesitancy from a nominal to a dichotomized (no/yes) categorical variable for primary analysis and used the Chi-squared test with Bonferroni correction for multiple comparisons to assess association of this outcome with characteristics of age, gender, race, political affiliation, and receipt of influenza in previous years. We then used a multivariable logistic regression to model the primary outcome variable with these same predictor characteristics.

To more intuitively depict COVID-19 vaccine acceptance (the converse of vaccine hesitancy), we chose the regression modelling technique of classification tree analysis and plotted results in a personograph. To prevent overfitting, we pruned the full tree to a smaller subtree using minimum-error pruning.

In terms of sample size calculation, we sought to power the primary outcome to 95% in assessment of its association with four characteristics - gender, race, age, and political affiliation. To adjust for multiple comparisons, we used an alpha level of 0.0125 (0.05/4) for statistical significance. Given the above information, the sample size needed to detect a small effect size (w) of 0.1 for a Chi-squared test with 1 degree of freedom [(2-1)*(2-1)] was 1,716.

We conformed our study reporting to the Strengthening the Reporting of Observational Studies in Epidemiology (STROBE) guidelines. We used JAMOVI v1.2.14.0 (Sydney, Australia) for statistical analyses.

## RESULTS

### Population Characteristics

Of 1,786 adult respondents, we excluded 30 for poor quality responses. Characteristics of the 1,756 respondents comprising the final study cohort are shown in Table 1. Most respondents self-identified as female (53%, n=931) and White (77%, n=1,356); their median age was 38 years (IQR 31-48). Approximately 85% (n=1,491) of the respondents had health insurance, and 78% (n=1,362) reported regular access to medical care. Most respondents lived with other people (84%, n=1474), including a significant other (71%, n=1,047), children <18 years of age (48%, n=706), and parents (21%, n=316). Approximately 8% (n=149) of respondents reported a previous diagnosis of COVID-19, and 20% (n=349) reported that one or more family members were previously diagnosed with COVID-19.

**Table 1.** Characteristics of U.S. Survey Respondents on Amazon Mechanical Turk (n=1,756).

| Characteristic | N (%) |
| --- | --- |
| **Gender** |  |
| Male | 810 (46) |

| **Characteristic** | **N (%)** |
| --- | --- |
| Female | 931 (53) |
| Non-binary | 15 (1) |
| **Age (median, IQR)** | 38 (31-48) |
| **Race/Ethnicity** |  |
| Non-Hispanic White | 1,356 (77) |
| Non-Hispanic Black | 152 (9) |
| Hispanic/Latinx | 100 (6) |
| Asian | 97 (6) |
| Other | 51 (3) |
| **<u>Covid-19 Diagnosis</u>** |  |
| Yes | 149 (8) |
| No / Unsure | 1,607 (91) |
| **Flu Vaccine Acceptance** |  |
| Yes | 1051 (60) |
| No | 675 (38) |
| Unsure | 30 (2) |
| **Education** |  |
| No High School | **3 (0)** |
| Grades 9-11 | 15 (1) |
| Grade 12 or GED | 180 (10) |
| College 1-3 years | 448 (26) |
| College 4 years or more | 799 (46) |
| Graduate or Professional degree | 311 (18) |
| **Region of Residence** |  |
| Northeast | 316 (18) |
| Midwest | 373 (21) |
| South | 681 (39) |
| West | 385 (21.9) |
| Unknown | 1 (0) |
| **Health Insurance** |  |
| Yes | 1,491 (85) |
| No | 219 (12) |
| Currently Applying for Insurance | 38 (2) |
| Unsure | 10 (1) |
| **Regular Access to Medical Care** |  |
| Yes | 1,361 (78) |
| No | 394 (22) |
| **Annual Income** |  |
| <u>Less than \$15k</u> | <u>90 (5)</u> |
| \$15k-25k | 178 (10) |
| \$26k-40k | 338 (19) |
| \$41k-59k | 381 (22) |
| \$60k-89k | 414 (24) |
| Greater than \$90k | 355 (20) |
| **<u>Living with Others</u>** |  |
| Yes | 1,474 (84) |
| No | 282 (16) |
| If yes, how many? (median, IQR) | 2 (1-3) |
| **Political Affiliation** |  |
| Conservative Republican | 252 (15) |
| Moderate Republican | 155 (9) |
| Liberal Republican | 115 (7) |
| Conservative Democrat | 127 (7) |
| Moderate Democrat | 236 (14) |
| Liberal Democrat | 458 (27) |
| Conservative Independent | 95 (6) |
| Moderate Independent | 109 (6) |
| Liberal Independent | 145 (8) |
| Unsure | 24 (1) |

IQR = interquartile range; k = 1000

### COVID-19 Vaccine Hesitancy

When asked about acceptance of the COVID-19 vaccine, 37.8% (663) were COVID-19 vaccine hesitant: 374 (21.3%) non-acceptors and 289 (16.5%) unsure about accepting. A similar proportion (40.1%, n=705) reported not receiving the influenza vaccine within the last five years.

In the multivariable logistic regression model (Table 2), respondents were more likely to be vaccine hesitant if they had not previously had an influenza vaccine (odds ratio [OR] 4.07, 95% confidence interval [CI] 3.26-5.07, p<0.01), identified as female (vs. male, OR 2.12, 95%CI 1.70-2.65, p<0.01), were Black (vs. White, OR 1.54, 95%CI 1.05-2.26, p=0.03), had a high school education or less (vs. college or more, OR 1.46, 95%CI 1.03-2.07, p=0.03), and were Republican (vs. Democrat, OR 2.41, 95%CI 1.88-3.10, p<0.01).

**Table 2.**
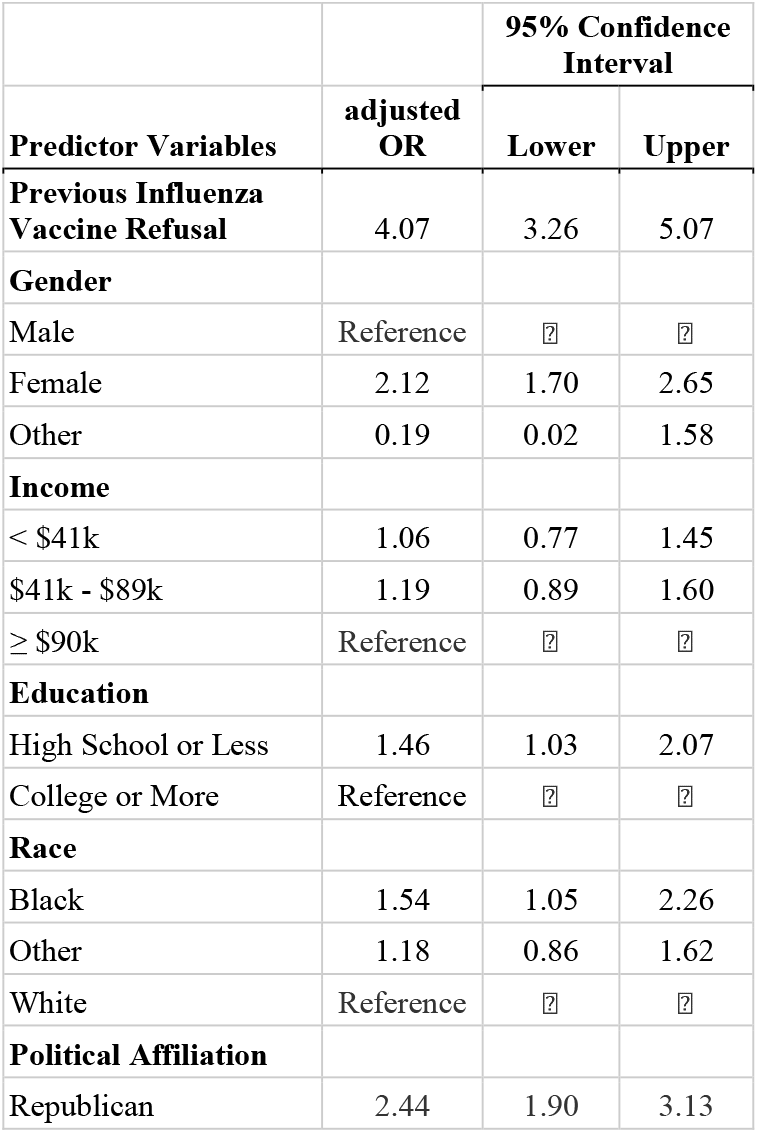
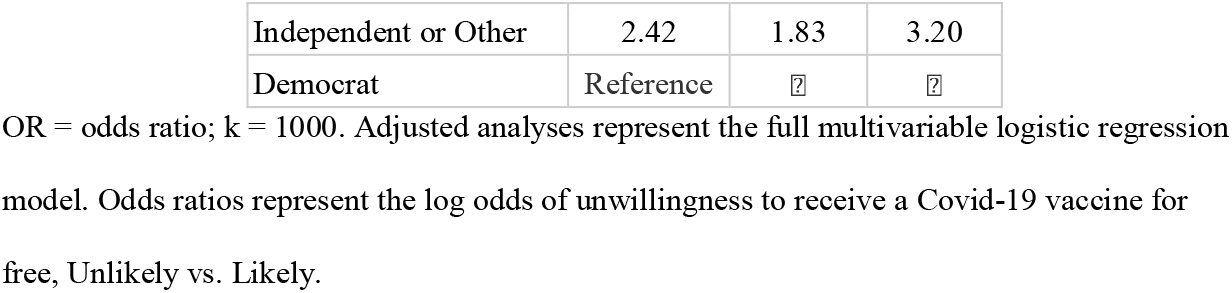
Predictors of Hesitancy of a Free Covid-19 Vaccine.

| Predictor Variables | adjusted OR | 95% Confidence Interval |  |
| --- | --- | --- | --- |
|  |  | Lower | Upper |
| **Previous Influenza Vaccine Refusal** | 4.07 | 3.26 | 5.07 |
| **Gender** |  |  |  |
| Male | Reference | ? | ? |
| Female | 2.12 | 1.70 | 2.65 |
| Other | 0.19 | 0.02 | 1.58 |
| **Income** |  |  |  |
| < \$41k | 1.06 | 0.77 | 1.45 |
| \$41k - \$89k | 1.19 | 0.89 | 1.60 |
| ≥ \$90k | Reference | ? | ? |
| **Education** |  |  |  |
| High School or Less | 1.46 | 1.03 | 2.07 |
| College or More | Reference | ? | ? |
| **Race** |  |  |  |
| Black | 1.54 | 1.05 | 2.26 |
| Other | 1.18 | 0.86 | 1.62 |
| White | Reference | ? | ? |
| **Political Affiliation** |  |  |  |
| Republican | 2.44 | 1.90 | 3.13 |
| Independent or Other | 2.42 | 1.83 | 3.20 |
| Democrat | Reference | ? | ? |

OR = odds ratio; k = 1000. Adjusted analyses represent the full multivariable logistic regression model. Odds ratios represent the log odds of unwillingness to receive a Covid-19 vaccine for free, Unlikely vs. Likely.

On classification tree analysis, previous receipt of an influenza vaccine and Democratic party political affiliation were significant predictors of COVID-19 vaccine acceptance (Fig 1).

**Fig 1.**
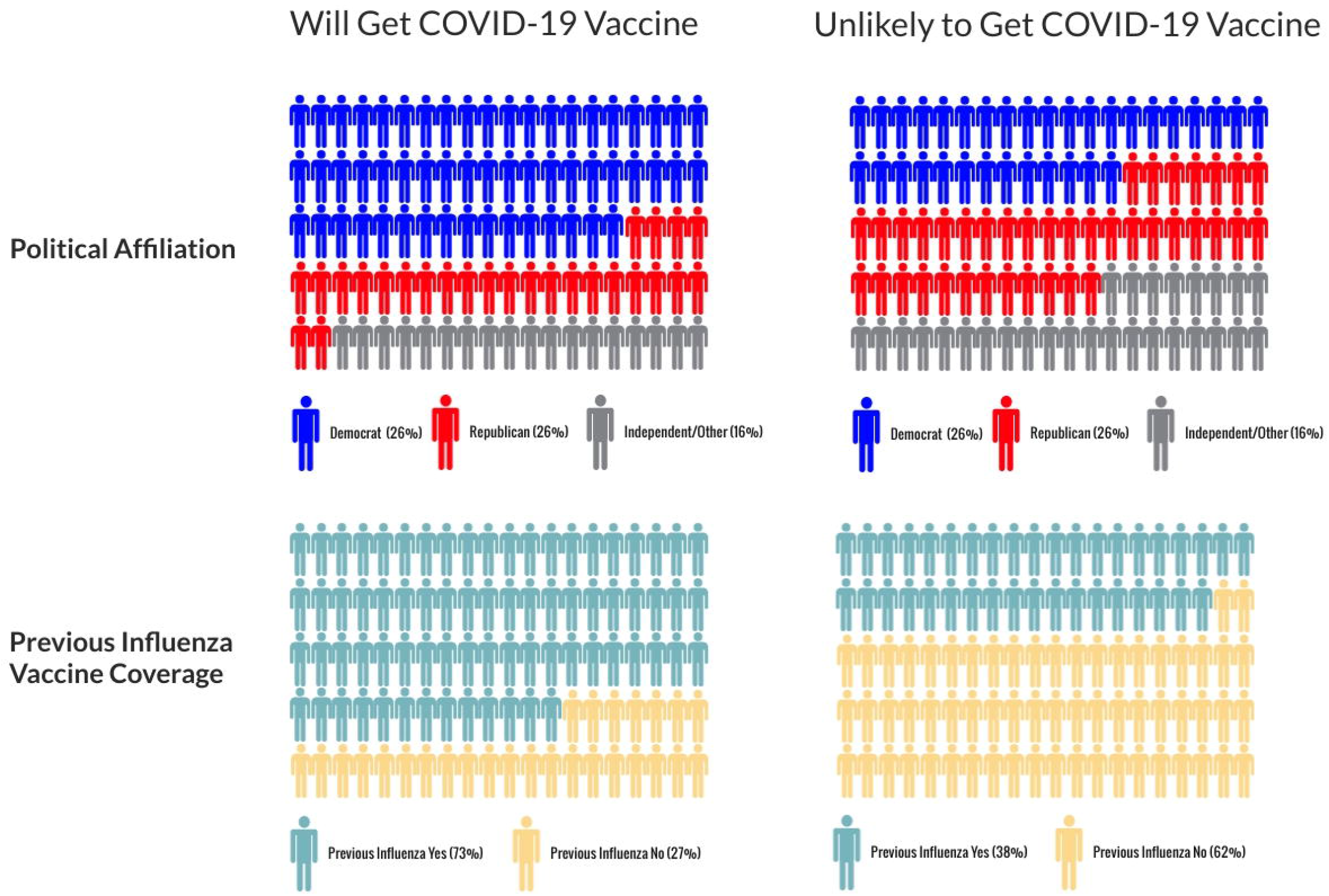
Main Predictors of Covid-19 Vaccine Hesitancy. Personograph plot of the classification tree analysis, which identified previous influenza vaccine coverage and political affiliation as significant predictors of COVID-19 vaccine hesitancy.

The main reasons given for vaccine were concerns about side effects and safety of the vaccine (75%, n=497), the need for more information about the vaccine (53%, n=351), and doubts regarding the efficacy of the vaccine (17%, n=110).

### Location of Receipt of COVID-19 Vaccination

Most vaccine acceptors (53%, n=576) preferred to receive it in their primary doctors’ offices or clinic. Pharmacies (32%, n=353) and dedicated vaccination locations (14%, n=154) were the next most preferred locations.

## DISCUSSION

Optimal health policy deliberations for COVID-19 vaccine distribution require consideration of vaccine hesitancy and reasons for refusal. We found significant vaccine hesitancy in the U.S. population that was more common in women, Blacks, and people with lower education levels or who identified as Republicans. Vaccine safety and side effects were the primary concerns, and over half of vaccine non-acceptors wanted more information before rendering a decision. Prior non-receipt of the influenza vaccine was the most powerful predictor of unwillingness to receive the COVID-19 vaccine. For those respondents willing to receive the COVID-19 vaccine, most indicated that they would prefer to receive it at their primary physician’s office/clinic.

Our data adds to the growing body of literature regarding vaccine hesitancy. A number of patient characteristics (socioeconomic status, level of education, health literacy, political affiliation, and race/ethnicity, among others), have historically played a role in attitudes toward vaccines (6,15,16). Beyond these patient level predictors, vaccine hesitancy also varies by vaccine type with childhood vaccines, such as MMR (measles, mumps, and rubella) and DTaP (diphtheria, tetanus, and pertussis), having much higher acceptance rates than adult vaccines (e.g., DTaP boosters, Pneumococcus, yearly influenza) (17).

Other unique characteristics of COVID-19 vaccine development may further complicate issues of vaccine acceptance. The unprecedented “warp” speed of research, development, and approval of the COVID-19 vaccines with significant public/governmental involvement and investment, has led some to speculate about their safety and efficacy (15). Disinformation and conspiracy theories about masks, transmission, therapeutics, and vaccines - amplified through social media and other venues - are also particularly vexing (18).

Driven in part by popular perception of poor efficacy and fear of side effects, influenza vaccine hesitancy is common (8–11,19). Given that influenza vaccine refusal appears to be predictive of hesitancy of COVID-19 vaccination, public health campaigns should emphasize the much higher efficacy of the COVID-19 vaccine (>90%) (20,21).

The racial/ethnic differences in vaccine hesitancy which we encountered in our study are highly concerning, but not unexpected given prior literature. Black and Hispanic/Latinx individuals have consistently lower influenza vaccination rates than their White counterparts (9– 11). Possible reasons for this historical difference include differences in racial consciousness leading to differential trust in the vaccine process and safety, general disparate trust in health care institutions, and limited knowledge of the specific vaccines (10,11). Unfortunately, vaccine hesitancy may further exacerbate the disproportionate effects of the pandemic on Latinx, African-American and Native American populations (22). Strategies to engage communities of color, including trusted messenger programs about safety of COVID-19 vaccines, will be essential to address this critical health disparity.

Previous research has shown associations between political affiliation and various health metrics and behaviors, including vaccination acceptance. Republican voters have been found to have lower self-reported influenza vaccination rates and an increased propensity for anti-vaccination beliefs when compared to Democrats (23,24). Our study corroborates these findings and suggests a need for political leaders of all parties to promote COVID-19 vaccination broadly among their constituencies.

Regarding our finding of association of lower education levels with vaccine hesitancy, prior literature has shown mixed results; some studies have found similar associations and others the opposite (6,11). Our results reaffirm the concept that information regarding vaccine safety and efficacy should be in language that is understandable by those with all levels of education.

One of the potential ways to address vaccine hesitancy is to ensure that vaccines are dispensed at locations where patients are most comfortable receiving them. Our results indicate that patients are most willing to go to their own clinics or physicians for vaccinations. Given that community pharmacies are embedded within neighborhoods and are seen as trusted sources for health information, they should also be prioritized for vaccine distribution (25,26).

Prior large studies have shown that the most effective efforts at reducing vaccine hesitancy are both multi-faceted and targeted at specific populations (7). A one-size-fits-all model is unlikely to work. Instead, a framework of engaging community and religious leaders, active messaging in various digital and non-digital media, education campaigns, targeted and incentivized vaccine drives, and wide distribution of vaccine at trusted sites will likely be required in order to decrease vaccine hesitancy.

The study has limitations. Responses were provided by interested survey respondents who had self-selected themselves into the Amazon Turk population (“Turkers”) who all had internet access, limiting the generalizability of our findings to other underserved populations. In this regard, Latinx respondents in this study were under-represented (6%) in comparison to their percentage in the general U.S. population (16.7%). Because the study platform does not allow for determination of how many people saw the invitation and did not participate, we could not calculate a true survey response rate.

In conclusion, COVID-19 vaccine hesitancy is common in the U.S. population and more prevalent in women, Blacks, people with lower education levels and Republicans. To improve efficient and equitable vaccine distribution, educational messaging campaigns should seek to address non-acceptors’ primary concerns of safety and side effects of the vaccine.

## Data Availability

De-identified data can be made upon request and with approval of the Mass General Brigham IRB.

